# Application and Reliability of Arm Use Intensities in the Free-Living Environment for Manual Wheelchair Users

**DOI:** 10.1101/2020.07.20.20157909

**Authors:** Brianna M. Goodwin, Omid Jahanian, Meegan G. Van Straaten, Emma Fortune, Stefan I. Madansingh, Beth A. Cloud-Biebl, Kristin D. Zhao, Melissa M. Morrow

**Affiliations:** Health Sciences Research and Kern Center for the Science of Health Care Delivery, Mayo Clinic Rochester, USA; Assistive and Restorative Technology Center, Physical Medicine and Rehabilitation Mayo Clinic Rochester, USA; Program in Physical Therapy, Mayo Clinic School of Health Sciences, Mayo Clinic, Rochester, USA

**Keywords:** spinal cord injury, inertial measurement units, wearable sensors, upper extremity

## Abstract

Arm use in individuals with spinal cord injury who use manual wheelchairs (MWC) is complex, characterized by a combination of overuse and a sedentary lifestyle. This study aimed to calculate arm use intensity levels for MWC users, describe the percentage of daily wear time MWC users and able-bodied individuals spend in each arm use intensity level, and test the reliabilities of the measurements for both MWC users and able-bodied individuals.

MWC users wore two inertial measurement units (IMUs) on their bilateral upper arms while performing six MWC-based activities in-lab. Video data were recorded and each second was coded as active or stationary. Acceleration-based signal magnitude area (SMA) ranges were defined for stationary, low, mid, and high arm use intensity levels. IMU data were also collected in the free-living environments for MWC users and able-bodied individuals for four days (3 weekdays and 1 weekend day). The SMA levels were applied to the free-living data from the dominant arm and the percentage of time spent in each level was calculated. The required number of days to achieve moderate, good, and excellent reliabilities was calculated.

Eight adult MWC users with SCI participated in the in-lab data collection and SMA arm use intensity levels were defined as, stationary: ⩽ 0.67g, low: 0.671 – 3.27g, mid: 3.271 – 5.87, and high: > 5.871. Six MWC users and 15 able-bodied individuals completed the free-living data collection. The dominant arm of both MWC users and able-bodied individuals was stationary for the majority of the day. The reliability analysis indicated that at least five and eight days of data are needed from MWC users and able-bodied individuals, respectively, to achieve reliable representation of their overall daily arm use intensities throughout a week.

Future research is needed to understand the recovery time associated with stationary arm use and if it differs between MWC users and matched able-bodied individuals. At least five days of data should be collected when utilizing these methods for MWC users. The methods presented here will contribute to understanding the mechanisms which cause increased shoulder pain and pathology for MWC users.

## I. Introduction

Almost 300,000 individuals in the United States have a spinal cord injury (SCI) with over 17,000 new injuries occurring each year [1]. The majority of individuals with SCI are non-ambulatory and require a wheelchair for their daily mobility [2]. Individuals with SCI who use a manual wheelchair (MWC users) have a high prevalence of musculoskeletal pain and injury [3, 4] and the shoulder is the most common site of this pain and pathology [5, 6]. Further, greater than 15% of individuals with acquired SCI reported shoulder pain as “unbearable” [3] and chronic MWC users are reported to be up to 10 times more likely to experience a rotator cuff tear compared to age-matched able-bodied individuals [4]. The upper extremity is essential for MWC users as it is used for both mobility and activities of daily living (ADLs). Monitoring how the arms are used in the free-living environment may lead to useful information about mechanisms of injury and different use patterns between MWC users and able-bodied individuals. These insights could lead to improved shoulder preservation guidelines for this population.

There are multiple contributing theories of shoulder pathology development for MWC users, including arm overuse and a sedentary lifestyle. Specifically, the load bearing nature of transfers and the repetitive task of propulsion are thought to lead to overuse of the arms for MWC users [7]. Overuse of the arms during MWC-based activities (mobility and ADLs) can contribute to increased pain and pathology in the shoulder for this population [8, 9]. However, the etiology of shoulder pain and pathology is multifactorial and is not due to arm overuse alone [10]. People with SCI are also overall more sedentary than able-bodied individuals, which may lead to a decrease in the overall use of the arms for MWC users compared to the able-bodied population [11, 12]. The contradiction of a more sedentary life-style and elements of arm overuse complicate the understanding of the link between the intensity of arm use of MWC users and the associated shoulder pain and pathology. In this paper, we define intensity of arm use as an acceleration-based measure of the magnitude of upper-arm movement calculated from the signal magnitude area (SMA). Intensity as described in this paper should not to be compared to intensity of physical activity related to energy expenditure or the rate of perceived exertion.

Wearable sensors such as accelerometers and inertial measurement units (IMUs) are low cost, accurate tools that have been used for monitoring and quantifying physical activity and clinical motion analysis in the able-bodied population and individuals with neurological conditions [13-15]. A few groups have created and validated overall energy expenditure or activity level thresholds specific for manual wheelchair users with wrist worn accelerometers [16, 17]. Additionally, metrics from accelerometers placed on the upper arm during propulsion [18] and a variety of other tasks [19] have been used to estimate physical activity levels. Although these methods are useful for estimating overall physical activity levels for individuals with spinal cord injury, they have not been used to understand the intensity of arm use, as a measure of overuse. Further, the reliability of these methods when applied to the free-living environment is largely unknown. The levels of activity and arm use vary from day to day within subjects due to environmental factors, individual characteristics, variations in daily schedules, and health condition. Therefore, it is required to investigate the number of days of monitoring that are needed to obtain a reliable representation of the overall arm use of a subject. A holistic daily view of the intensity of arm use MWC users exhibit compared to able-bodied individuals may uncover patterns which provide context to understanding the increase in pain and pathology for MWC users.

This study aimed to define and explore arm use intensity levels (stationary, low, mid, and high), estimate the percentage of daily time MWC users and able-bodied individuals spend in each arm use intensity level throughout a typical day, and estimate the reliability of the metrics for both cohorts.

## II. Methods

### A. Study Design

Two separate data collections were employed to (1) define the arm use intensity levels and (2) estimate the percentage of daily time MWC users and able-bodied individuals spend in each arm use intensity level and test the reliabilities (Fig. 1). First, in-lab IMU and video data, previously collected from a sample of MWC users [20], were utilized to determine the range of SMA magnitudes that defined each of the arm use intensity levels (stationary, low, mid, and high). After classification accuracy was assessed for the stationary and active threshold, the in-lab levels were applied to IMU data collected in the free-living environment from a different sample of MWC users and able-bodied individuals. The percentage of time spent in each arm use intensity level was calculated. Additionally, the single-day reliabilities of all metrics were calculated and used to estimate the required number of monitoring days needed to achieve a reliable representation of daily arm use intensity levels for both cohorts.

**Fig. 1.**
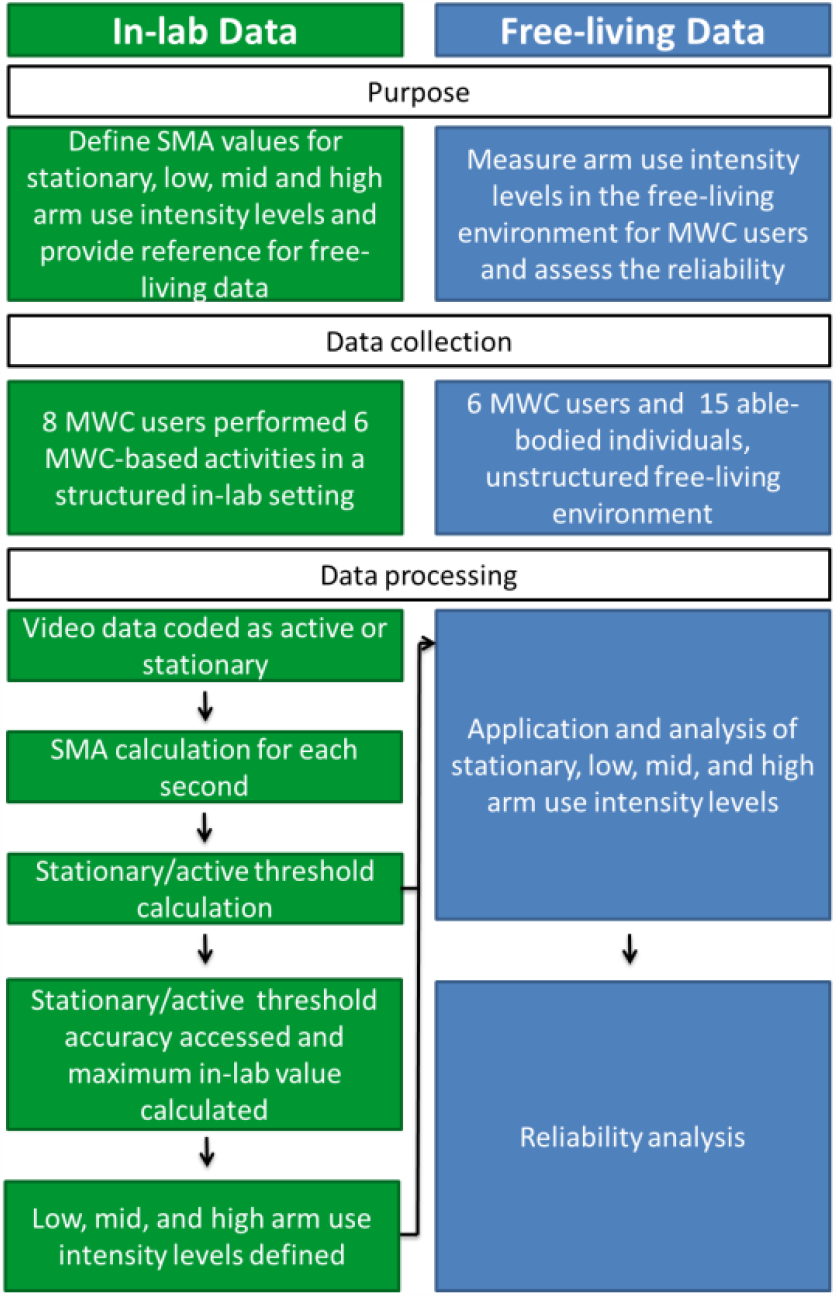
The purpose, data collection, and data processing flow of defining signal magnitude area (SMA) arm use intensity levels for the in-lab and free-living data utilized in this study.

All aspects of the study were approved by Mayo Clinic Institutional Review Board. Individuals with SCI who were 18-70 years of age and using a MWC as their main mode of mobility for at least one year were recruited for the in-lab and free-living field data collections through querying medical databases and care providers of local clinics. In-lab inclusion criteria included active shoulder range of motion needed to complete the lab tasks (∼150° of humeral elevation). Able-bodied individuals age 18-70 were recruited to participate in the free-living data collections; they were not age and sex-matched to the MWC users. Participants were excluded in all parts of the study if they had prior significant surgery or injury to the shoulder or had cognitive impairments which may have limited their ability to follow instructions and adhere to the protocol. Additionally, able-bodied individuals and MWC users were excluded from the free-living data collections if they had previous diagnosis of a complete supraspinatus tear or were unable to receive an MRI of one or both shoulders as the data presented here is part of a larger longitudinal study following the natural history of the rotator cuff.

### B. In-lab Data Collection for Intensity Level Calculation

IMU and video data were collected while participants performed six MWC-based activities as part of an ancillary study [20, 21]. Two IMUs were secured with straps to each of the participant’s upper arms (Emerald, APDM, Inc., Portland, OR) and data were collected at 128 Hz. Video data were collected at 60 Hz using a handheld digital camera throughout the entire data collection. Participants were asked to complete six MWC-based activities: 1) counter height reaching (36 inches above the ground), 2) overhead reaching (54 inches above the ground), 3) 6.8 kg cross-body lifting of a backpack from the floor on the side of his/her wheelchair to a plinth on the other side, 4) level transfers between the MWC and plinth, 5) level MWC propulsion (on rollers), and 6) 5° incline MWC propulsion (on rollers with a wooden board under the casters to create an incline). During the reaching tasks, each participant retrieved and returned an aluminum (soup) can (0.45 kg) from a table (counter height) or shelf (overhead). For reaching and cross-body lifting, each participant used the arm that they self-selected as their trailing limb during their daily car transfers into the car or the side to which they transfer most frequently. Additionally, participants self-selected their cadence during propulsion.

Participants were asked to perform 10 reaches for both counter height and overhead reaching, three cross body lifts (from ground to plinth and back to ground), and six transfers (from MWC to plinth or plinth to MWC). They were additionally asked to perform two MWC propulsion trials: propulsion on level rollers for approximately two minutes and propulsion on a simulated incline condition for approximately 15 seconds. In cases where technical difficulties occurred, additional trials were performed, if possible. In some cases, fewer numbers of trials or shorter duration of MWC propulsion were performed based on the participant’s physical capacity. On average participants completed 11 ± 3 counter height reaches, 11 ± 3 overhead reaches, 4 ± 2 cross-body lifts, 6 ± 2 transfers, 110 ± 51 seconds of level propulsion, and 25 ± 5 seconds of inclined propulsion. The activities and time naturally occurring between them were included in the data used to define arm use intensity levels to ensure inclusion of natural resting periods.

### C. Free-living Data Collection

Similar to the in-lab data collection, participants were fit with two IMUs on their bilateral arms (Emerald or Opal, APDM, Inc., Portland, OR). Participants were asked to wear the sensors for at least eight hours on four consecutive days (one weekend day and three weekdays), charge the sensors each night and not change their regular activities. Each participant was provided in-person, written, and video instructions to increase protocol adherence. Participants were provided a pre-paid envelope or met study staff in person to return the sensors after completion of the data collection.

### D. Data Processing

Raw acceleration data from the IMUs were downloaded through Motion Studio (APDM, Inc., Portland, OR) for both the in-lab and free-living data collections. The linear acceleration data were used to quantify the intensity of arm use by calculating the SMA with a custom MATLAB (Mathworks, Natick, MA) code. SMA is a measure of the intensity of movement over one second; the methods are described elsewhere [22]. In short, the acceleration signal was filtered with a centered median filter to reduce noise spikes (window size of 3 frames) [22, 23]. The gravitational component of the signal was then calculated by using a third-order zero phase lag elliptical low pass filter with 0.25 Hz cut-off frequency, 0.01 dB passband ripple and -100 dB stopband ripple. The gravitational component of the acceleration signal was subtracted from the original signal to leave the gravitational component due to body movement and the SMA was calculated for each second of data [22].

### E. Arm Use Intensity Level Defintions (In-lab Data)

VLC media player (VideoLAN Organization, Paris, France) was used to view all in-lab video data for analyses. For the six in-lab MWC-based activities and the time naturally occurring between them, one rater (EF) with more than eight years of movement and video analysis experience coded each second of data as either stationary or active [20]. A threshold between stationary and active arm use was calculated from a receiver operating characteristic (ROC) curve; the SMA value which maximized specificity and sensitivity for a subset of data (6 of 8 participants, ∼78% of total data) was chosen as the threshold (0.67 g). The remaining data (2 of 8 participants, ∼22% of total data) were lumped together and used assess the accuracy of this threshold.

The low, mid, and high arm use levels were then defined. The maximum in-lab SMA values from all participants were averaged (8.46 ± 1.59 g). For the dominant arm, the maximum SMA was reached during incline propulsion for four participants and during transfers for four participants. Three evenly spaced intensity intervals were then defined between the maximum value of active movement and the minimum value of active movement (active/stationary threshold), yielding low, mid, and high intensity levels. SMA analyses of the active portions of the in-lab MWC-based activities provide a reference for the free-living SMA data. Therefore, the mean, maximum and minimum active SMA values were calculated for each of the six in-lab MWC-based activities for each participant.

### F. Arm Use Intensity Level Application to Free-living Data

Free-living data were excluded if less than eight hours of useable data were collected for each day (after elimination of non-wear time). All data were visually inspected to ensure non-wear time was eliminated. Eight hours was chosen as the minimum collection period for a full day as a balance between sensor battery life and inclusion of data. Utilizing the arm use intensity levels from the in-lab data, the daily percentages of time that individuals spent in each intensity level were calculated for each participant’s dominant arm. The average time across all four days of data was calculated for both cohorts.

### G. Reliability Statistical Analysis

Single-day reliabilities of arm use intensity levels were calculated based on the weekday & weekend (4 days: one weekend and three weekdays) measurements and only weekday (3 weekdays) measurements, using reliability analyses in SPSS 25 (IBM Corp., Armonk, NY). When available weekday & weekend measurements were consecutive days from Sunday through Wednesday and only weekday measurements were consecutive days from Monday through Wednesday. Single-day reliability was defined as single measure Intraclass Correlation Coefficients (ICC), calculated based on one-way random effects model [15, 24]. For both cohorts the required days of monitoring needed to achieve moderate, good, and excellent reliabilities were calculated using the Spearman-Brown prophecy formula based on the weekday and weekday & weekend measurements separately [25]. Reliability coefficient values between 0.5 and 0.75 are considered as moderate, 0.75 and 0.9 as good, and higher than 0.9 as excellent reliability [26]. For all analyses, statistical significance was set at an alpha level of p < 0.05.

## III. Results

### A. Arm Use Intensity Levels During In-lab MWC-based Activities

Eight participants were included in the in-lab data collection (Table I). Six of the eight participants were used to calculate the threshold between stationary and active arm use; the remaining two participants were used to assess the accuracy of the threshold. The active/stationary threshold had specificity, sensitivity, and overall accuracy values of 94%, 80%, and 89%, respectively. The SMA ranges for each level are shown in Table II. Representative data from one participant completing all activities are shown in Fig. 2. The mean, maximum and minimum SMA for each activity give context to the arm use intensity levels participants utilized to accomplish MWC-based activities in the lab (Table III).

**TABLE I.** In-lab and free-living (Manaual Wheelchair (MWC) and able-bodied individuals) participant demographics

|  | In-Lab | MWC | Able-bodied |
| --- | --- | --- | --- |
| **Number of participants** | 8 | 6 | 15 |
| **Age (years), mean + SD (range)** | 39.4 ± 12.6<br>(23-61) | 51.5 ± 9.6<br>(31-59) | 42.2 ± 12.0<br>(25-59) |
| **Sex** | 7 males/1 female | 4 males/2 females | 12males/3 females |
| **Injury Level** |  |  |  |
| C6-C7 | 4 | 1 | - |
| T1-T8 | 4 | 1 | - |
| T9-L1 | 0 | 4 | - |
| **Time since injury (years), mean ± SD (range)** | 9.6 ± 11.3<br>(1-33) | 21.9 ± 13.1<br>(2-35) | - |
| **Number of participants who reported presence of pain** | 5 | 5 | 2 |

**TABLE II.** The defined in-lab signal magnitude area (sma) range for each arm use intensity level and the mean ± standard devaition percentage of the day the domiannt arm of the manaual wheelchair users (mwc) and able-bodied individuals sepnt in each arm use level in the free-living environment

| Arm use intensity level | SMA Range (g) | MWC (% of day) | Able-bodied (% of day) |
| --- | --- | --- | --- |
| **Stationary** | SMA ≤ 0.67 | 65.6 ± 11.4 | 59.6 ± 12.6 |
| **Low intensity** | 0.67 < SMA ≤ 3.27 | 29.7 ± 9.9 | 29.7 ± 8.9 |
| **Mid intensity** | 3.27 < SMA ≤ 5.87 | 4.0 ± 1.8 | 9.4 ± 5.2 |
| **High intensity** | SMA > 5.87 | 0.7 ± 0.6 | 1.2 ± 1.3 |

**TABLE III.** The mean ± standard devaition of the minimum, maximum, and mean signal magnitude area (sma) for the eight participants who completed the six in-lab manual wheelchair-based activities

|  |  | Minimum SMA (g) | Maximum SMA (g) | Mean SMA (g) |
| --- | --- | --- | --- | --- |
| **Counter height reaching** | Reaching arm | 0.5 ± 0.2 | 2.6 ± 0.4 | 1.2 ± 0.3 |
|  | Non-reaching arm | 0.2 ± 0.2 | 2.0 ± 1.0 | 0.7 ± 0.5 |
| **Overhead reaching** | Reaching arm | 0.6 ± 0.2 | 4.1 ± 0.7 | 2.0 ± 0.2 |
|  | Non-reaching arm | 0.2 ± 0.1 | 3.4 ± 1.8 | 1.1 ± 0.5 |
| **Cross-body lifts** | Dominant | 0.3 ± 0.2 | 5.0 ± 0.9 | 1.7 ± 0.3 |
|  | Non-Dominant | 0.3 ± 0.1 | 4.8 ± 0.9 | 1.6 ± 0.2 |
| **Transfers** | Dominant | 0.3 ± 0.2 | 7.5 ± 1.2 | 2.3 ± 0.4 |
|  | Non-Dominant | 0.2 ± 0.1 | 8.1 ± 1.7 | 2.2 ± 0.4 |
| **Level propulsion** | Dominant | 1.0 ± 0.3 | 7.1 ± 1.4 | 5.4 ± 1.2 |
|  | Non-Dominant | 1.0 ± 0.4 | 6.8 ± 2.0 | 5.0 ± 1.4 |
| **Incline propulsion** | Dominant | 1.0 ± 0.5 | 7.4 ± 1.6 | 5.3 ± 1.0 |
|  | Non-Dominant | 1.0 ± 0.6 | 7.1 ± 1.9 | 5.0 ± 1.1 |

**Fig. 2.**
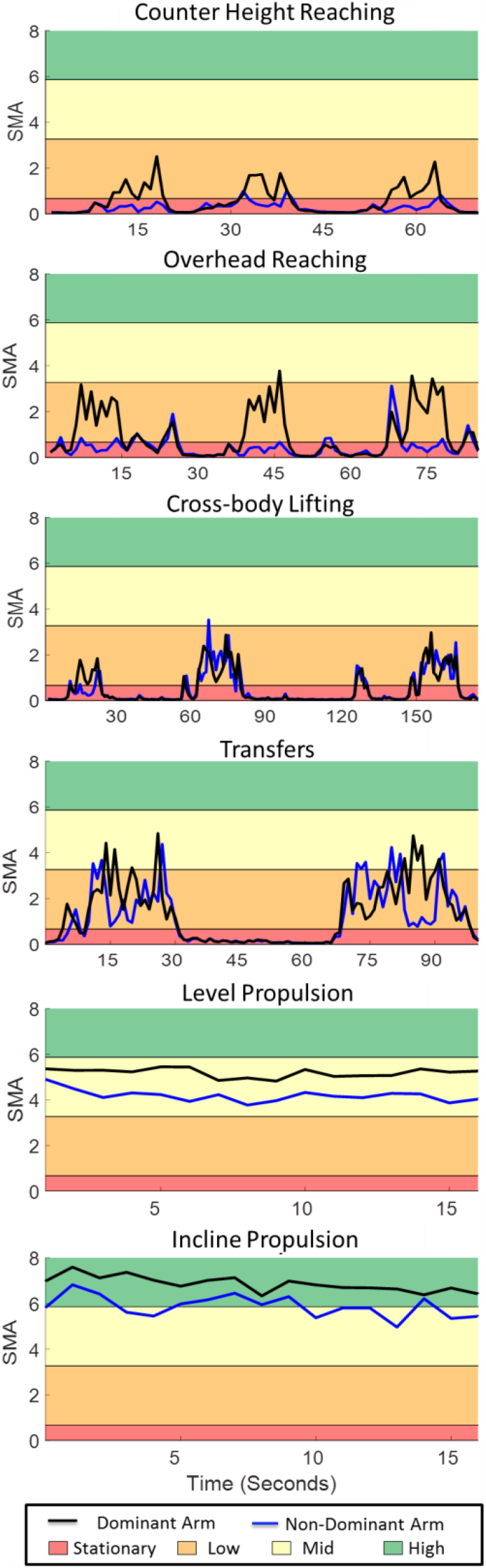
Representative signal magnitude area (SMA) data from one participant completing three counter hieght reaches, three over head reaches, threee cross body lifts, two transfers, level propolsion and incline propolsion in a structured lab setting for their dominant (black line) and non-dominant (blue line). The shaded areas represent the am use intensity levels (red: stationary, orange: low, yellow: mid, greeen: high).

### B. Percentage of Free-living Datain Arm Use Levels

Six MWC users and 15 able-bodied individuals were enrolled in the free-living data collection (Table I). Data were collected on average for 9.7 ± 0.8 and 10.3 ± 1.2 hours for the MWC and able-bodied cohorts, respectively. Both cohorts spent the majority of their day stationary (Table II). The MWC and able-bodied cohorts spent less than 5% (∼30 minutes) and 11% (∼66 minutes) of the day in and above mid arm use intensity levels, respectively.

### C. Relability of Arm Use Levels

All MWC users and 73% (11/15) of able-bodied individuals had consecutive weekday & weekend measurements from Sunday to Wednesday and only weekday measurements from Monday to Wednesday. When consecutive days could not be achieved (4/15 able-bodied individuals), data from three non-consecutive weekdays and one weekend day were used in analysis. Single-day reliabilities of arm use intensity levels ranged from moderate to poor for both MWC and able-bodied cohorts (Table IV). The results from the weekday & weekend measurements indicated that a good reliability (>0.75) is reached when monitoring MWC users for five days and able-bodied individuals for eight days. The results from only weekday measurements indicated that a good reliability (>0.75) is reached when monitoring MWC users for three weekdays and able-bodied individuals for four weekdays (Table 4).

**TABLE IV.** Intraclass reliability coefficients (iccs) for mwc and able-bodied cohorts based on only weekday measurements (3 days) and weekday & weekend (3 weekdays, 1 weekend) measurements and the number of required days of monitoring to obtain moderate, good, and excellent reliabilities

|  | Single day reliability | F Test | Days of monitoring to achieve reliabilities of: |  |  |
| --- | --- | --- | --- | --- | --- |
|  | ICCs | Sig | 0.5 | 0.75 | 0.9 |
| **MWC Cohort – Only Weekdays** |  |  |  |  |  |
| **Stationary** | 0.620 | 0.006 | 0.61 | 1.84 | 5.52 |
| **Low** | 0.587 | 0.009 | 0.70 | 2.11 | 6.33 |
| **Mid** | 0.454 | 0.035 | 1.20 | 3.61 | 10.82 |
| **High** | 0.487 | 0.026 | 1.05 | 3.16 | 9.48 |
| **MWC Cohort – Weekdays & Weekend** |  |  |  |  |  |
| **Stationary** | 0.441 | 0.011 | 1.27 | 3.80 | 11.41 |
| **Low** | 0.413 | 0.016 | 1.42 | 4.26 | 12.79 |
| **Mid** | 0.389 | 0.021 | 1.57 | 4.71 | 14.14 |
| **High** | 0.444 | 0.011 | 1.25 | 3.76 | 11.27 |
| **Able-bodied Cohort – Only Weekdays** |  |  |  |  |  |
| **Stationary** | 0.638 | 0.001 | 0.57 | 1.70 | 5.11 |
| **Low** | 0.722 | 0.001 | 0.39 | 1.16 | 3.47 |
| **Mid** | 0.591 | 0.001 | 0.69 | 2.08 | 6.23 |
| **High** | 0.567 | 0.001 | 0.76 | 2.29 | 6.87 |
| **Able-bodied Cohort – Weekdays & Weekend** |  |  |  |  |  |
| **Stationary** | 0.543 | 0.001 | 0.84 | 2.52 | 7.57 |
| **Low** | 0.567 | 0.001 | 0.76 | 2.29 | 6.87 |
| **Mid** | 0.292 | 0.007 | 2.42 | 7.27 | 21.82 |
| **High** | 0.646 | 0.001 | 0.55 | 1.64 | 4.93 |

## IV. Discussion

The primary purpose of this study was to explore the application of arm use intensity levels (stationary, low, mid, and high) and estimate the percentage of time, the dominant arm of MWC user and able-bodied individuals, spend in each arm use intensity level. We also aimed to calculate the single-day reliability of the metrics and estimate the number of days which are required to obtain a reliable representation of overall daily arm use intensities. The results indicated the dominant arm of both cohorts was stationary and in low arm use intensity levels for the majority of the day. To achieve good reliabilities for all arm use intensity levels throughout an entire week (weekdays and weekend days) at least five and eight days of data are needed for MWC users and able-bodied individuals, respectively.

Although no tasks were identified in the free-living environment in this study, the in-lab MWC-based activities help give context to the measured arm use intensity levels during free-living. For example, the data presented here suggests that level and incline propulsion occurs in the mid and high arm use levels. Additionally, other in-lab wheelchair-based tasks achieve multiple arm use intensity levels; for instance, during transfers arm use intensities range from stationary to high levels. It is critical to recognize that the arm use metrics are only based off acceleration values and no load is accounted for. The load bearing nature of transfers is thought to contribute to arm overuse for MWC users and lead to increased pathology in this population [7]. Therefore, further research is needed to determine if SMA values alone are associated with pain and pathology of the shoulder.

The able-bodied individuals trended toward spending a larger percentage of time in the mid arm use intensity level than MWC users; however due to the exploratory nature of this study and non-matched cohorts, no statistical analyses were performed to test for significance. Based on the lab data, level propulsion, inclined propulsion, transfers, overhead reaching, and cross body lifting had portions of the activity that achieved a mid intensity level. If lab-based arm use intensities are comparable to free-living arm use intensities, we may conjecture that the able-bodied cohort performs activities in the mid arm use intensity level more frequently than MWC users. However, we did not collect reference activities for the able-bodied cohort, so the comparison across intensities is meant as an initial description into characterizing how MWC users and able-bodied adults may differ in how they use their arms throughout a typical day. Additionally, this study was not designed to compare across groups, as noted in the differences between cohort size and age. This study is part of a larger longitudinal study investigating the natural history of rotator cuff disease. Future analyses will use matched MWC and able-bodied pairs to determine if differences are significant and clinically meaningful.

The MWC and able-bodied cohorts spent most of their day with their arms stationary. Although, some of this time likely includes rest and recovery of the musculoskeletal system of the arms, likely some of this time does not allow for recovery. Industrial ergonomic studies have shown that short periods of rests (less than five seconds) do not provide an adequate recovery time during repetitive upper extremity tasks [27, 28]. The SMA was defined in one second epochs; therefore, it is possible that active arm use occurred immediately before and after stationary seconds. Further research is needed to build on the current methodologies to incorporate the duration of stationary levels and evaluate the difference in these stationary levels between MWC users and able-bodied individuals. Additionally, ergonomics literature has reported that for every 10 minutes of work a 90 second rest is required [29]. Future work is also needed to investigate if MWC users achieve the recommended rest to work ratios and to determine if they correlate to pathology development among MWC users.

The results from the reliability analysis indicate that at least five and eight days of data are needed from the MWC users and able-bodied individuals, respectively, to achieve reliable representation of their overall daily arm use intensity throughout a week (weekdays and weekends). However, only four and three days of data are needed to obtain a reliability representation of only weekdays for MWC users and able-bodied individuals, respectively. The difference between the weekday & weekend and only weekday analysis is primarily due to a difference in arm use intensity levels during weekdays versus weekends. One other study has investigated the reliability of wheelchair-based metrics from wrist-worn accelerometery data for individuals with SCI who use MWCs [24]. The metrics included measures of physical activity intensity, wheeling quantity, and movement quality from data collected during in- and out-patient settings. Their results suggest that four days of data are needed for good reliability of movement quality and three days are required for most other metrics in the out-patient setting [24]. Reference [24] found no difference between weekdays and weekends in wheelchair distance traveled. This difference between the current study and [24] could be in part due to a difference in metrics measured, time since injury, or other differences in study design. The results presented here included individuals on average 20+ years since injury, while the cohort in [24] included participants 300-400 days post injury. Further research is needed to understand if weekday and weekend differences shown in the current study occur due to differing arm use intensity patterns, but similar propulsion levels.

It is important to understand the study limitations when interpreting our findings. First, no angular kinematic data or information about when the arm was loaded (push phase of propulsion, lifting an object, transfers, etc.) were included in these analyses. Understanding both the load and arm position during each arm use intensity level would allow more insight in pathomechanisms of injury for MWC users and how it differs from able-bodied individuals. Implementing algorithms which calculate and integrate arm loading and the position of the arm with the arm use intensity levels were out of the scope of the current study; however, analysis of the arm position during arm use intensity levels is an active area of research for our group. Additionally, the arm use intensity levels were defined based on a small cohort of MWC users and only six in-lab activities. A larger cohort of MWC users may perform activities differently and activities may be performed at higher SMA levels in the free-living environment. Further, able-bodied individuals may perform ADLs with different arm use intensity levels than the MWC cohort, however the use of a single set of levels allowed for description and descriptive comparison between both cohorts. Only six MWC-based activities were performed in the in-lab data collection and therefore other activities which the MWC and able-bodied cohorts perform throughout a day will occur in various intensities of arm use. Finally, the data presented in this study is from a small non-matched cohort of MWC users and able-bodied controls. Caution should be taken when drawing conclusions from differences between the arm use intensity levels of the MWC and able-bodied cohorts. Ultimately our research group aims to develop metrics from wearable technologies which inform the risk of shoulder pathology development or progression and/or development of pain.

This study aimed to define arm use intensity levels, apply the levels to free-living data of MWC users and able-bodied individuals, and measure the number of days of data needed to achieve a reliable representation of arm use. The results of this study suggest that differences between arm use intensity levels of MWC users and able-bodied individuals warrant further study. Additionally, five and eight days of data are needed to achieve good reliabilities for all arm use levels throughout weekdays and weekends for MWC users and able-bodied individuals, respectively. Understanding the intensity levels of arm use for MWC users compared with able-bodied individuals may aid in understanding the mechanisms of rotator cuff pathology which occur more frequently in MWC users than able-bodied individuals. The causes of increased shoulder pain and pathology for MWC users are multifactorial and the intensities of arm use are only one piece of a complicated puzzle. Other characteristics of use, such as duration of arm use bouts, amount of rest/recovery between repetitive motions, over-head humeral elevation and loading contribute to pain and pathology. Future work from our group is focused on understanding factors of arm overuse during the daily lives of MWC users and the relationship between overuse and the progression of shoulder pain and pathology.

## Data Availability

Data is available upon request.

## Acknowledgment

This publication was made possible by funding from the National Institutes of Health (grant no. R01 HD84423-01), Craig H. Neilsen Foundation for Spinal Cord Injury Care and Research Honoring Robert D. Brown Jr, Mayo Clinic Robert D. and Patricia E. Kern Center for the Science of Health Care Delivery, and the National Center for Advancing Translational Sciences (UL1 TR002377).

## Notes

### Competing Interest Statement

The authors have declared no competing interest.

### Author Declarations

All aspects of the study were approved by Mayo Clinic Institutional Review Board.

